# Prevalence and Predictors of Depression, Anxiety and Stress among Elderly in the aftermath of COVID-19: A Quantitative Study from Central India

**DOI:** 10.1101/2022.03.10.22272024

**Authors:** Varun Malhotra, Danish Javed, Rajay Bharshankar, Vijender Singh, Namita Gautam, Shweta Mishra, Digpal S. Chundawat, Anuradha Kushwah, Gyanendra Singh

**Affiliations:** Department of Physiology, All India Institute of Medical Sciences, Bhopal, M.P., India; Department of AYUSH, All India Institute of Medical Sciences, Bhopal, M.P., India; Department of Psychiatry, All India Institute of Medical Sciences, Bhopal, M.P., India; Department of Community Medicine, All India Institute of Medical Sciences, Bhopal, M.P., India; Department of Nursing, All India Institute of Medical Sciences, Bhopal, M.P., India

**Author notes:** **Correspondence address:** Prof. Dr. Vijender Singh, Professor & Head, Department of Psychiatry, AIIMS, Saket nagar, Bhopal, M.P., India-462020.

**Keywords:** Aged, Frail Elderly, Geriatric Psychiatry, Depression, Anxiety, Loneliness, Home Isolation, COVID-19, Pandemics, Mental Health

## Abstract

**Background:** Elderly persons have been more prone to depression, anxiety and stress during COVID-19 pandemic. They need more care and support towards mental health during these difficult times.

**Methodology:** This was a cross-sectional and quantitative study conducted in the state of Madhya Pradesh, during the month of March 2021 to August 2021. Participants were recruited from a population aged more than 60 years, those who were able to read and write Hindi/English or having at least one family member; reporting to a tertiary care teaching hospital during the second wave of COVID-19 in India. Those who were confirmed COVID-19 cases & undergoing treatment, with diagnosed mental health disorders and who didn’t give consent were excluded. A semi-structured questionnaire along with DASS-21 scale was completed by participants online.

**Results:** Out of 690 subjects, 7.25% had mild and moderate depression, 0.58% had severe and extremely severe depression. Mild and moderate anxiety symptoms were reported by 9.56%, 2.46% reported severe and extremely severe anxiety. Mild and moderate stress was reported by 4.78%, while 0.42% were severely and extremely stressed. A positive statistical association was found between alcohol addiction and depression (*p*=0.028). The elderly subjects had a nap the day time were significantly less depressed during COVID-19 pandemic (*p*=0.033). It was found that older the subjects, more were they anxious during the pandemic (*p*=0.042). An association was found between alcohol addiction and stress (*p*=0.043).

**Conclusions:** Depressive symptoms in participants were positively correlated with alcohol addiction. Females reported higher level of stress. There is a felt need to formulate psychological interventions for elderly to improve their mental health and psychological resilience. We need to tackle and fight the stigma, fear and anxiety related to the COVID-19, which is greater than disease itself.

**Key message:** *What is already known:* Psychological problems like anxiety, depressive symptoms, fearfulness, a state of uncertainty and stress are common in all age groups; furthermore older adults are more prone to develop mental health issues in wake of stressful situations.

*What this study adds to:* About one fourth of elderly developed anxiety, depression and stress during the COVID-19 pandemic.

*Effect on practice and policy:* There is a need for proactive identification through screening of elderly for mental health issues following unprecedented stress like COVID-19 pandemic.

## INTRODUCTION

On 31^st^ December 2019, a novel viral pneumonia originating from Wuhan, China was reported to the World Health Organization.[1] Changes to daily life have been swift and unprecedented, as cases of the virus surged, the death toll escalated, and draconian measures to contain the spread of the disease increased across regions of the globe. Although there has been substantial attention towards measures to identify people with the corona virus infection, identifying the mental health care needs of people impacted by this pandemic had been relatively neglected.[2] For instance, in a recent, large survey of people highly susceptible to the corona virus infection, the prevalence rate of traumatic stress was at an alarming rate (73.4%), depression (50.7%), generalized anxiety (44.7%), and insomnia (36.1%).[3] Although these findings are disturbing, but they are not isolated, as research on the psychological impact of previous global disease outbreaks has demonstrated clear links between pandemic-related anxiety and elevated symptoms of stress, anxiety, contamination concerns, health anxiety, post-traumatic stress, and suicidality.[4,5]

In fact, the finding that those with a positive COVID-19 status reported significantly elevated levels of corona virus anxiety score as compare to non-infected and the importance of assessing and treating the psychological needs of those infected with the virus.[6] If some expert opinions are considered, then that would mean that up to 70% of the world’s population could potentially need both medical and psychological care following COVID-19 infections.[7]

During the initial phase of COVID-19 outbreak in China, more than half of the respondents rated their psychological impact as moderate-to-severe, and about one-third reported moderate-to-severe anxiety. Specific, updated and accurate health related information and certain precautionary measures such as covering mouth when coughing and sneezing, washing hands with soap immediately after coughing, sneezing, rubbing the nose, or after touching contaminated objects, and wearing a mask and social distancing were associated with a lower psychological impact of the outbreak and lower levels of stress, anxiety, and depression.[8]

The governments also provided mental health services through different channels including hotline, online consultation, online courses and outpatient consultation, but more attention should be paid to depression and anxiety.[9] There have been increased worries and apprehensions among the elderly regarding acquiring the COVID-19 infection. Elderly persons have higher perceived needs to deal with their mental health difficulties. The older adults were confronted with challenges like non-availability of attendants to help them with tasks of daily living, difficulty in accessing medical help, and perceived apprehension of existing morbidities. The older adults do not have an easy access to the mental health professionals. They are vulnerable to feelings of loneliness and helplessness. They fear that even if they survive the pandemic, social order would change. Some fear worsening of morbidity and deterioration in quality of life. In the general community, overall wellbeing, activity levels, and sleep were reported to be affected along with associated changes in cognitive functioning.[10]

Stress during an infectious disease outbreak can include fear and worry about, health of loved ones that manifests as changes in sleep or eating patterns, difficulty in sleeping or concentrating worsening of chronic health problems, worsening of mental health conditions, and increased use of alcohol, tobacco, or other drugs. Elderly are more prone to depression, anxiety and stress.

This study was planned to find out the prevalence of depression, anxiety and stress during COVID-19 times and their association with demographic and clinical factors and is aimed to fill these very lacunae in the available research especially in elderly population. It is important to study the mental health impacts in elderly for planning effective intervention strategies for this population.[11]

## MATERIAL AND METHODS

### Study Design

A quantitative cross sectional study was conducted with duration of 06 months from March 2021 to August 2021 in a tertiary care teaching hospital in the state of Madhya Pradesh, in central India. The study population consisted of elderly persons (≥60 years of age) who were free of any COVID-19 related symptoms at the time of data collection. The subjects reported their mental health experiences of first as well as second wave of COVID-19 in India.

### Sample selection

A sample size of 690 subjects was recruited through online survey after the approval of Institutional Human Ethics Committee of All India Institute of Medical Sciences, Bhopal, Madhya Pradesh, India (Letter No. IHEC-LOP/2020/EF0221, dated: 24/12/2020). A written consent was taken from the participants after explaining the detail about research work. The sample size for this study was calculated as per a previous study by Gorrochategi et al. using DASS-21 scale, [12] taking prevalence of depression and anxiety in the age group ≥ 60 years as 18.6% and 13.5% respectively and applying the formula for cross-sectional study as 4pq/d2, taking 80% power, relative precision to be 20%, and non-response to be 10%, it was found that minimum sample size required was 482 and 690 respectively. The data collection was done through a semi-structured questionnaire used along with DASS-21 scale through a google form to recruit the participants.

### Inclusion criteria

Subjects of age ≥ 60 years of age and residing in the state of Madhya Pradesh; those who were able to read and write Hindi/English or having at least one family member who could read and write Hindi/English so that they could answer the questions of DASS-21 and semi structured questionnaire and they were be free from any COVID-19 related symptoms at the time of data collection were recruited for this study.

### Exclusion criteria

Those who were confirmed COVID-19 positive cases, with ongoing treatment at the time of study, people with diagnosed mental health disorders and those who did not give consent were excluded.

## RESULTS

Out of 690 subjects on applying DASS-21, 28(4.06%) suffered from mild depression, 22 (3.19%) moderate depression, 2(0.29%) severe depression and 2(0.29%) from extremely severe depression. 21(3.04%) were mildly anxious, 45(6.52%) had moderate anxiety, 6(0.87%) severe anxiety and 11(1.59%) extremely severe anxiety. 19(2.75%) elderly subjects were stressed mildly, 14(2.03%) were moderately stressed, 1(0.14%) severely stressed and 2(0.28%) extremely stressed. (**Table-1**)

**Table 1:** Socio-demographic data of the sample of Elderly.

| Variables |  |  |  |  |  |  |  |
| --- | --- | --- | --- | --- | --- | --- | --- |
| Age (n= 690) (years) | Mean = 67.45 (SD ± 6.49) | 95% CI= 66.96 to 67.93 | (60-92) Median=66 | Mean age (Male=67.77 ± 6.52) | Mean age (Female= 66.47 ± 6.28) | P value= 0.0227* ANOVA |  |
| Range (age) | 60 - <65 | 65 - <70 | 70 - <75 | 75 - <80 | 80 - <85 | 85 - <90 | 90 - <95 |
| N (%) | 273(39.57%) | 169(24.49%) | 141(20.43%) | 69(10.00%) | 29(4.20%) | 7(1.01%) | 2(0.29%) |
| Sex | Male | Female |  |  |  |  |  |
| N (%) | 517(74.93%) | 173(25.07%) |  |  |  |  |  |
| Location | Bhopal | Outside Bhopal |  |  | Urban | Rural |  |
| N (%) | 683(98.99%) | 7(1.01%) |  |  | 684(99.13%) | 6(0.87%) |  |
| Marital Status | Married | Widowed/widower | Unmarried |  |  |  |  |
| N (%) | 638(92.46%) | 48(6.96%) | 4(0.58%) |  |  |  |  |
| Education | Uneducated | Primary education | Matric/Intermediate | Graduate | Post-graduate & above |  |  |
| N (%) | 18(2.61%) | 40(5.80%) | 158(22.90%) | 240(34.78%) | 234(33.91%) |  |  |
| Employment status | Retired | Government organization |  | Business | Private job | House wife |  |
| N (%) | 529(76.67%) | 50(7.24%) |  | 43(6.23%) | 34(4.93%) | 34(4.93%) |  |
| Healthcare worker | No | Yes, but not working during pandemic |  |  | Frontline workers | Second line workers |  |
| N (%) | 665(96.38%) | 16(2.13%) |  |  | 7(1.01%) | 2(0.29%) |  |
| Depression | Normal | Mild | Moderate | Severe | Extremely Severe |  |  |
| (N, %) | 636(92.17%) | 28(4.06%) | 22(3.19%) | 2(0.29%) | 2(0.29%) |  |  |
| (Mean Score, ±SD, 95%CI) | 1.16±2.15 (1.00 to 1.33) | 10.79±0.99 (10.40 to 11.17) | 16.36±2.19 (15.39 to 17.34) | 22.00±0.00 (22.00 to 22.00) | 37.00±7.07 (-26.53 to 100.53) |  |  |
| Anxiety | Normal | Mild | Moderate | Severe | Extremely Severe |  |  |
| (N, %) | 607(87.97%) | 21(3.04%) | 45(6.52%) | 6(0.87%) | 11(1.59%) |  |  |
| (Mean Score, ±SD, 95%CI) | 1.03±1.77 (0.89 to 1.17) | 8.00±0.00 (8.00 to 8.00) | 11.64±1.67 (11.14 to 12.15) | 17.00±1.10 (15.85 to 18.15) | 23.27±6.53 (18.89 to 27.66) |  |  |
| Stress | Normal | Mild | Moderate | Severe | Extremely Severe |  |  |
| (N, %) | 654(94.78%) | 19(2.75%) | 14(2.03%) | 1(0.14%) | 2(0.28%) |  |  |
| (Mean Score, ±SD, 95%CI) | 2.41±3.80 (2.12 to 2.70) | 16.53±0.90 (16.09 to 16.96) | 21.57±1.79 (20.54 to 22.60) | 30.00± 0.00 (30.00to 30.00) | 38.00±5.66 (-12.82 to 88.82) |  |  |

Among the participants, COVID-19 information level was very high (91%). Around 19% participants reported that their work was adversely affected during to COVID-19 pandemic. Fear of infection was testified either ‘occasionally’ or ‘many times’ by 39% participants. However, their self-assessment for physical health and mental health was reported as ‘good’ and ‘very good’ in 86% and 87% respectively. Around 70% elderly conveyed that they were spending more than one hour of time in watching television or social media. **(Table-2)**

**Table 2:** Prevalence of conditions/symptoms across socio-demographic variables (n = 690).

| Variables |  |  |  |  |  |
| --- | --- | --- | --- | --- | --- |
| A. Tobacco addiction | Never | Occasionally | Regularly |  |  |
|  | 560(81.16%) | 102(14.78%) | 28(4.06%) |  |  |
| B. Alcohol addiction | Never | Occasionally | Regularly |  |  |
|  | 568(82.32%) | 116(16.81%) | 6(0.87%) |  |  |
| C. COVID-19 status | COVID-19 positive | Negative, but tested for COVID-19 | Corona Recovered | Knowing someone affected by virus | None |
|  | 4(0.58%) | 51(7.39%) | 27(3.91%) | 32(4.64%) | 576(83.48%) |
| D. COVID-19 Information level | Large extent | Some extent | Minimum | No information |  |
|  | 502(72.75%) | 127(18.41%) | 45(6.52%) | 16(2.32%) |  |
| E. Financial condition affected in pandemic | No effect | Minimum | Moderately | Very much |  |
|  | 327(47.39%) | 110(15.94%) | 160(23.19%) | 93(13.48%) |  |
| F. Work adversely affected during COVID-19 | Yes | No |  |  |  |
|  | 130(18.84%) | 560(81.16%) |  |  |  |
| G. Boredom/frustration during lockdown | Never | Occasionally | Many times |  |  |
|  | 395(57.25%) | 238(34.49%) | 57(8.26%) |  |  |
| H. Fear of infection | Never | Occasionally | Many times |  |  |
|  | 397(60.98%) | 214(32.87%) | 40(6.14%) |  |  |
| I. Stress due to insufficient supply | Never | Occasionally | Many times |  |  |
|  | 501(72.61%) | 173(25.07%) | 16(2.32%) |  |  |
| J. Travel abroad since January 2020 | Yes | No |  |  |  |
|  | 25(3.62%) | 665(96.38%) |  |  |  |
| K. Physical health (Self-assessment) | Very good | Good | Average | Below average |  |
|  | 310(44.92%) | 286(41.45%) | 86(12.46%) | 8(1.16%) |  |
| L. Mental health (Self-assessment) | Very good | Good | Average | Below average |  |
|  | 352(51.01%) | 252(36.52%) | 78(11.3%) | 8(1.16%) |  |
| M. Any mental health issue | Yes | No |  |  |  |
|  | 31(4.49%) | 659(95.51%) |  |  |  |
| N. Sleep quality | Poor | Average | Satisfactory |  |  |
|  | 17(2.46%) | 155(22.46%) | 518(75.07%) |  |  |
| O. Sleep disturbances | Never | occasionally | 1-2 times per week | Daily |  |
|  | 358 (51.88%) | 285 (41.30%) | 24(3.48%) | 23(3.33%) |  |
| P. Time to fall asleep | Within 30 minutes | 30 minutes to 1 hour | More than an hour |  |  |
|  | 533 (77.23%) | 121(17.54%) | 36(5.22%) |  |  |
| Q. Difficulty to get back to sleep again at night | Sometimes difficult | No difficulty | Very much |  |  |
|  | 211(30.58%) | 448(64.93%) | 31(4.49%) |  |  |
| R. Early morning awakening even if don't want | Never | Sometimes | 1-2 times per week | Almost daily |  |
|  | 399(57.74%) | 167(24.17%) | 15(2.17%) | 109(15.77%) |  |
| S. Feeling not refreshed in morning | Never | Mostly | 1-2 times per week | Almost daily |  |
|  | 477(69.13%) | 115(16.67%) | 70(10.14%) | 28(4.06%) |  |
| T. Sleep in daytime | Never | Occasionally | 1-2 hours daily | 2-3 hours daily |  |
|  | 187(27.10%) | 210(30.43%) | 250(36.23%) | 43(6.23%) |  |
| U. Change in you food intake pattern | No | Occasionally | 1-2 days per week | Daily |  |
|  | 590(85.51%) | 84(12.17%) | 6(0.87%) | 10(1.45%) |  |
| V. Regular bowel and bladder movements | Yes | Occasional disturbances | 1-2 days per week | More than these |  |
|  | 505(73.19%) | 170(24.64%) | 7(1.01%) | 8(1.16%) |  |

| W. | Time spend on the Internet and TV per day | Less than 1 hour | 1-2 hours | 2-4 hours | More than 4 hours |
| --- | --- | --- | --- | --- | --- |
|  |  | 202(29.27%) | 253(36.67%) | 162(23.48%) | 73(10.58%) |

On linear and multiple regression, a positive significant statistical association was found between alcohol addiction and depression (*p*=0.028). (**Table -3**)

**Table 3:**
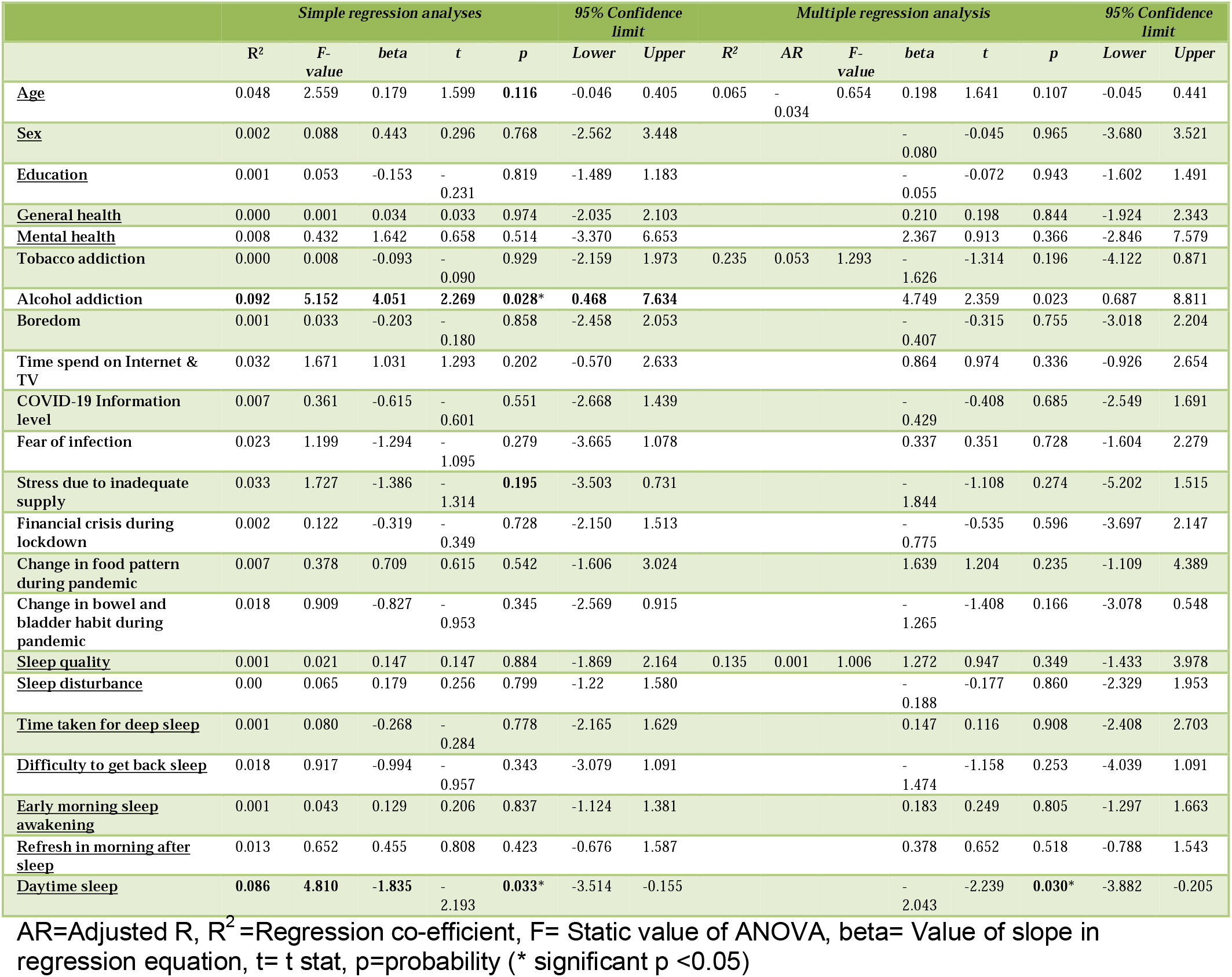
Results for Regression analysis with DASS-D score computed as dependent variable (n = 54).

The elderly subjects who had more day time sleeping hours were significantly less depressed in these times of COVID-19 (*p*=0.033). (**Table -3**)

A positive significant association was found for age of participants and levels of anxiety. The older the subjects and more were they anxious during the pandemic (*p*=0.042). (**Table-4**)

**Table 4:** Results for Regression analysis with DASS-A score computed as dependent variable (n = 83).

|  | Simple regression analyses |  |  |  |  | 95% Confidence limit |  | Multiple regression analysis |  |  |  |  | 95% Confidence limit |  |  |
| --- | --- | --- | --- | --- | --- | --- | --- | --- | --- | --- | --- | --- | --- | --- | --- |
|  | R<sup>2</sup> | F-value | beta | t | p | Lower | Upper | R<sup>2</sup> | AR | F-value | beta | t | p | Lower | Upper |
| <u>Age</u> | **0.049** | **4.256** | **0.188** | **2.063** | **0.042\*** | 0.007 | 0.369 | **0.088** | **0.028** | **1.478** | **0.179** | 1.904 | **0.061\*** | -0.008 | 0.366 |
| <u>Sex</u> | 0.029 | 2.389 | 1.929 | 1.546 | 0.126 | -0.554 | 4.411 |  |  |  | 1.866 | 1.358 | 0.178 | -0.869 | 4.602 |
| <u>Education</u> | 0.008 | 0.631 | -0.440 | -0.794 | 0.429 | -1.543 | 0.663 |  |  |  | -0.039 | -0.064 | 0.949 | -1.249 | 1.172 |
| <u>General health</u> | 0.001 | 0.059 | 0.198 | 0.243 | 0.809 | -1.422 | 1.817 |  |  |  | -0.151 | -0.151 | 0.880 | -2.135 | 1.833 |
| <u>Mental health</u> | 0.007 | 0.559 | 0.558 | 0.748 | 0.457 | -0.926 | 2.043 |  |  |  | 1.004 | 1.104 | 0.273 | -0.807 | 2.814 |
| <u>Tobacco addiction</u> | 0.007 | 0.605 | 0.773 | 0.778 | 0.439 | -1.205 | 2.752 | 0.063 | -0.067 | 0.484 | -0.231 | -0.192 | 0.848 | -2.627 | 2.165 |
| <u>Alcohol addiction</u> | 0.035 | 2.959 | 2.386 | 1.720 | **0.089** | -0.373 | 5.145 |  |  |  | 2.084 | 1.192 | 0.237 | -1.400 | 5.567 |
| <u>Boredom</u> | 0.015 | 1.229 | 0.999 | 1.109 | 0.271 | -0.795 | 2.795 |  |  |  | 0.554 | 0.528 | 0.599 | -1.538 | 2.645 |
| <u>Time spend on Internet & TV</u> | 0.002 | 0.189 | 0.284 | 0.435 | 0.665 | -1.016 | 1.584 |  |  |  | -0.059 | -0.080 | 0.936 | -1.530 | 1.412 |
| <u>COVID-19 Information level</u> | 0.003 | 0.276 | -0.452 | -0.525 | 0.601 | -2.163 | 1.259 |  |  |  | 0.166 | 0.173 | 0.863 | -1.739 | 2.070 |
| <u>Fear of infection</u> | 0.009 | 0.700 | 0.799 | 0.837 | 0.405 | -1.102 | 2.702 |  |  |  | 0.096 | 0.136 | 0.892 | -1.317 | 1.510 |
| <u>Stress due to inadequate supply</u> | 0.004 | 0.295 | 0.498 | 0.543 | 0.589 | -1.328 | 2.325 |  |  |  | -0.096 | -0.074 | 0.941 | -2.655 | 2.464 |
| <u>Financial crisis during lockdown</u> | 0.002 | 0.141 | 0.239 | 0.376 | 0.708 | -1.028 | 1.507 |  |  |  | 0.031 | 0.028 | 0.978 | -2.138 | 2.199 |
| <u>Change in food pattern during pandemic</u> | 0.029 | 2.447 | 1.259 | 1.564 | **0.122** | -0.343 | 2.862 |  |  |  | 1.078 | 1.142 | 0.257 | -0.803 | 2.959 |
| <u>Change in bowel and bladder habit during pandemic</u> | 0.002 | 0.132 | -0.326 | -0.363 | 0.718 | -2.115 | 1.463 |  |  |  | -0.583 | -0.599 | 0.551 | -2.523 | 1.357 |
| <u>Sleep quality</u> | 0.002 | 0.158 | -0.356 | -0.397 | 0.692 | -2.141 | 1.429 | 0.055 | -0.035 | 0.614 | -1.020 | -0.913 | 0.364 | -3.247 | 1.207 |
| <u>Sleep disturbance</u> | 0.006 | 0.465 | 0.456 | 0.682 | 0.497 | -0.874 | 1.785 |  |  |  | 0.758 | 0.825 | 0.412 | -1.073 | 2.589 |
| <u>Time taken for deep sleep</u> | 0.003 | 0.205 | 0.374 | 0.453 | 0.652 | -1.269 | 2.018 |  |  |  | 0.594 | 0.543 | 0.589 | -1.587 | 2.775 |
| <u>Difficulty to get back sleep</u> | 0.001 | 0.026 | -0.143 | -0.161 | 0.872 | -1.904 | 1.618 |  |  |  | -0.211 | -0.193 | 0.848 | -2.389 | 1.968 |
| <u>Early morning sleep awakening</u> | 0.000 | 0.000 | 0.003 | 0.005 | 0.996 | -0.977 | 0.982 |  |  |  | -0.211 | -0.379 | 0.705 | -1.315 | 0.894 |
| <u>Refresh in morning after sleep</u> | 0.004 | 0.359 | 0.277 | 0.599 | 0.550 | -0.641 | 1.195 |  |  |  | 0.186 | 0.379 | 0.705 | -0.791 | 1.163 |
| <u>Daytime sleep</u> | 0.033 | 2.735 | 1.039 | 1.654 | **0.102** | -0.212 | 2.291 |  |  |  | 1.059 | 1.579 | **0.119** | -0.277 | 2.395 |

AR=Adjusted R, R<sup>2</sup>=Regression co-efficient, F= Static value of ANOVA, beta= Value of slope in regression equation, t= t stat, p=probability (\* significant p <0.05)

On multiple regression analysis, a positive statistical association was found between alcohol addiction and stress (*p*=0.043); females were more stressed as compared to males (*p*=0.045) and people who were more educated were more stressed during the pandemic (*p*=0.036). (**Table-5**)

**Table 5:**
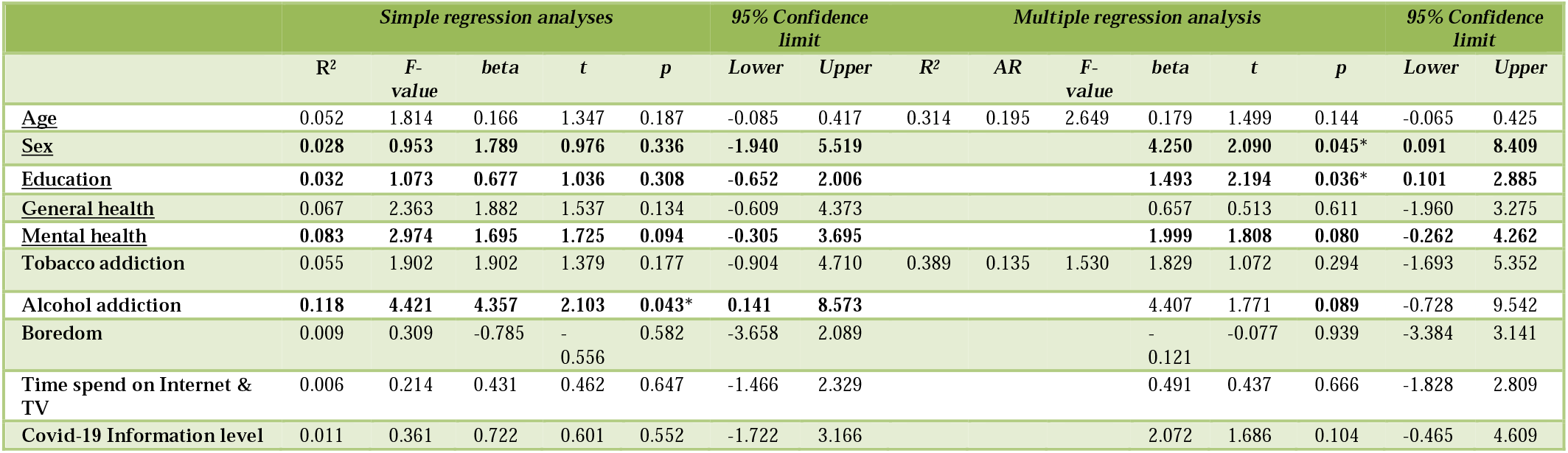
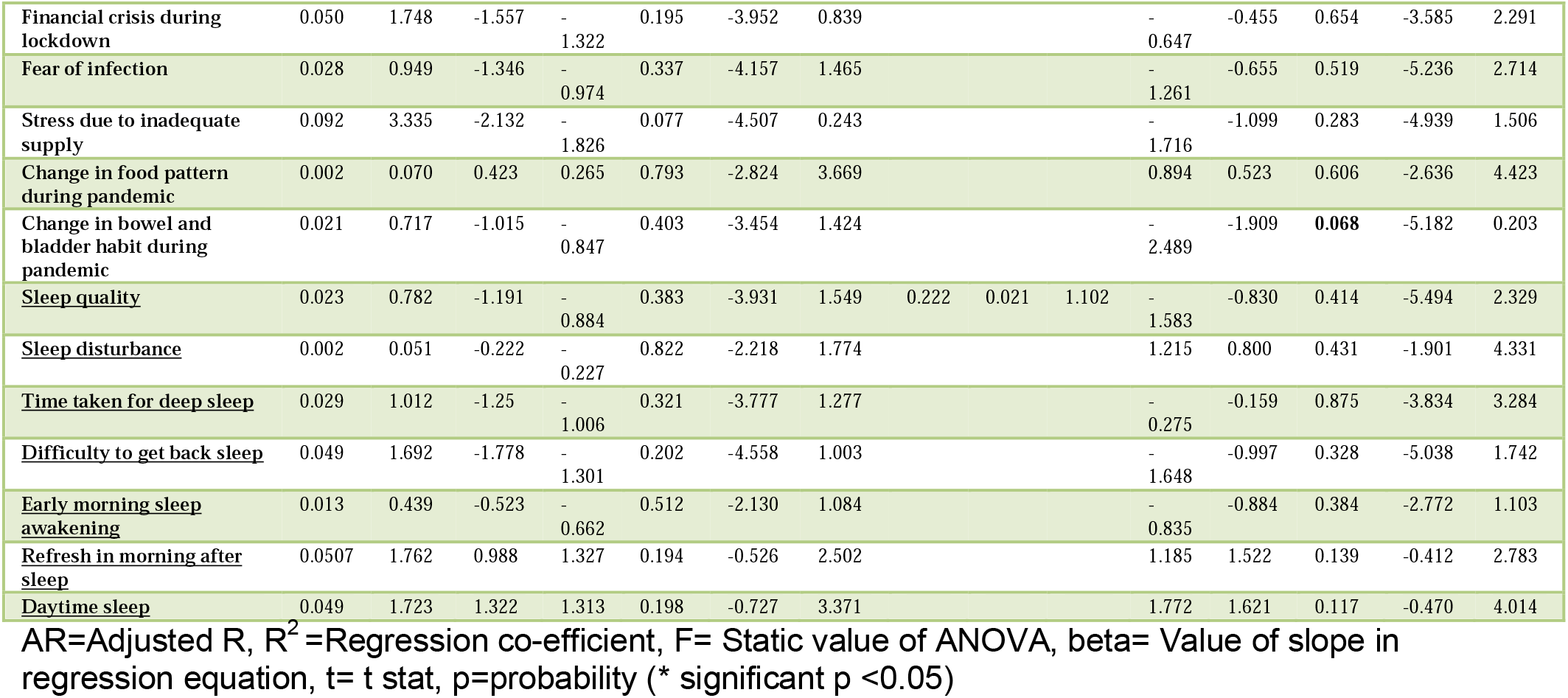
Results for Regression analysis with DASS-S score computed as dependent variable (n = 36).

## DISCUSSION

This study involved quantitative cross sectional assessment of elderly on online platform utilizing DASS-21 local language (Hindi) alongside English. The sampling method included a convenience sample of 690 respondents.

We found a point prevalence of 7.25% for mild and moderate depression and 0.58% for severe and extremely severe depression; 9.56% for mild and moderate anxiety and 2.46% for severe and extremely severe anxiety; 4.78% for mild and moderate stress and 0.42% for severe and extremely stress.

The older adults have a higher risk for severe respiratory problems due to COVID-19 including the risk for hospitalization, the requirement of ventilatory support and a high mortality rate contributing to significant stress, anxiety, and other related mental health problems. They are likely to experience more stress and difficulty in accessing essential services due to the restrictions enforced to maintain the social distancing to prevent the spread of COVID-19. Additionally, they are less likely to have social contact using technology and social media which are useful for maintaining social contact despite the need for physical distancing. The resulting isolation is one of the important risk factors for mental health problems like depression and anxiety in older adults.[13] All these factors may explain the higher prevalence of depression, anxiety and stress in around 25% of sample in our study.

It was found that 4.49% elderly were already having some pre-existing mental health issues (Table-2). According to the Centre for Disease Control and Prevention (CDC), older adults with COVID-19 are more likely to be hospitalized (31-59%) and die of it (4-11%). They also have higher chances of worsening of pre-existing medical and psychiatric illnesses because of difficulty in ensuring regular medication and routine health care due to COVID-19 related restrictions.[14]

### Mental health issues

Alcohol addiction, time spent on internet and television, higher COVID-19 information level, more fear of COVID-19 infection, higher stress due to inadequate supply, financial crisis during lockdown, absurd change in food and bowel pattern during pandemic, sleep disturbance in form of long time taken for deep sleep, difficulty to get back to sleep, early morning sleep awakening, non-refreshing sleep and daytime sleepiness were the factors found to be related with the mental health of elderly population (Table-2).

Literature suggests that, sleeplessness, feeling anxious, boredom, panic attacks, nightmares, feeling of emptiness, fear of contracting COVID-19, fear of spreading the infection to others, health anxiety, feeling of imprisonment, anxiety related to uncertainty about future, anxiety about death and dying in unnatural circumstances without access to other relatives are some of the psychological issues that can occur in older adults. Some of them may develop depression, anxiety, hypochondriasis, post-traumatic stress disorder, substance abuse/withdrawal in these stressful situations. Those living in old age homes/residential set-ups or living alone are the most vulnerable during this pandemic in terms of mental health.[15]

Findings of our study showed that the COVID-19 pandemic had worsened depressive mood and apathy among the community-dwelling older adults. The results were remarkable especially in elderly aged less than 75 years. Higher physical activity is important in maintaining mental health.[16] Individuals aged more than 75 years might be less affected by COVID-19 because their baseline physical activity was lesser than that of individuals aged less than 75 years. Additionally, this study suggests that elderly aged less than 75 years, who are usually considered to be relatively robust, however they are at high risk for worsened depressive mood and apathy. It was noted that mental abnormalities in this situation may have long lasting effects like post-traumatic stress disorder and has brought forth the risk of the current lack of mental health screening systems.

Additionally, apart from the physical effects of COVID-19, significant psychological effects such as anxiety, depression, and loneliness are shown to affect individuals of all ages including the older adult population; individuals aged 65 years and older.[17] Social isolation and lack of social contacts related to the COVID-19 pandemic potentially worsens anxious and depressive feelings, increasing the risk for adverse health outcomes in the high-risk older adult population.[18] One systematic review focusing on data of last 15 years of prevalence of depression in older adults reported rates of major depression ranging broadly from 0.9% to 9.4%.[19] Specific factors in older adults, including somatic illness, poor health, cognitive or functional impairment, history of depression, lack of close social contacts are identified as main predictors of depressive disorders.[20] In a systematic review of 11 cross-sectional and longitudinal studies conducted during this pandemic, a strong association was identified between psychological effects such as worry, anxiety, and mental anguish with pandemic behaviors such as social distancing and enhanced hygiene measures such as frequent hand washing.[21] The literature identifies feelings of isolation, loneliness, anxiety, and depression are prevalent in the older adult population, which were potentially worsened during a pandemic.[22]

### Associated Factors

In our study, it was found that older people who slept during the day were substantially less depressed. (*p*=0.033) **(Table 3)**. As per literature, napping was found to improve cognitive and emotional health in older persons.[23] A mid-day nap is shown to be advantageous in healthy, young people. A nap in the middle of the day reduces drowsiness while improving executive function, memory consolidation and subsequent learning.[24-26]

### Recommendations for intervention

The first step is increasing the awareness about mental health issues among the older adults and their family members via social media (Online programs, website, online forum, group email or messages). Utilizing the services of community health workers or trained social workers for the screening of older adults at old age homes/assisted living facilities for mental health issues. Telemedicine can be used for consultations requiring more detailed psychiatric evaluation and prescription of medication. Provision of mental health services to older adults in China during the COVID-19 pandemic was found to be challenging, with the “digital divide,” quarantines, and “restrictions on movement” having significant impact.[27] Responses to COVID-19 for older adults can borrow general mental health recommendations from historic pandemic influenza preparedness initiatives including a focus on strengthening social networks and community support, communication networks, directive leadership and stress management. One of the few positives in the pandemic literature in older adults is the possibility that older adults may experience more adaptive coping in a crisis than younger adults, as suggested by a study in Hong Kong during the SARS epidemic.[28] This form of resilience may relate to their lived experience of coping with adversity and could potentially be harnessed now.

Lessons learnt from earlier pandemics like SARS have proved that regular telephonic contact, monitoring and contact with family, relevant and updated information, caring for the general medical and psychological needs, and respecting their personal space and dignity are important components of mental health care in the elderly.[29]

Similar to other situations leading to any disaster, most of the older adults are likely to have sub-syndromal mental health issues like anxiety and depressive symptoms related to the threat of COVID-19. This will require brief psychological and psychosocial intervention that can be delivered by any health care personnel, volunteers, with some guidance and training from mental health professionals. Older adults need reassurance that most of the mental health issues experienced in these situations are normal reactions to abnormal stress. Yoga, meditation, healthy diet, mental stimulation through home-based activities with appropriate safety precautions is essential. Brief relaxation exercises and supportive therapy can be done for those having severe psychological distress. It is warranted to establish a system that could promptly screen and support the mental health of older adults and other mentally vulnerable populations.

## CONCLUSION

Based on a cross-sectional quantitative study, it was found that a significant proportion of elderly had depression (7-8%) anxiety (9-10%) and stress (5%) making it around 1/4^th^ of the sample of 690 respondents. Alcohol related problems were associated with stress & depression and age of subjects was associated with level of anxiety in linear fashion. Day time nap was found to have a positive effect on the mental health of participants. Based on the findings of this study it may be considered necessary to screen elderly population for mental health issues and provide timely interventions for the same.

## Data Availability

All data produced in the present study are available upon reasonable request to the authors

---

**Figure 1:**
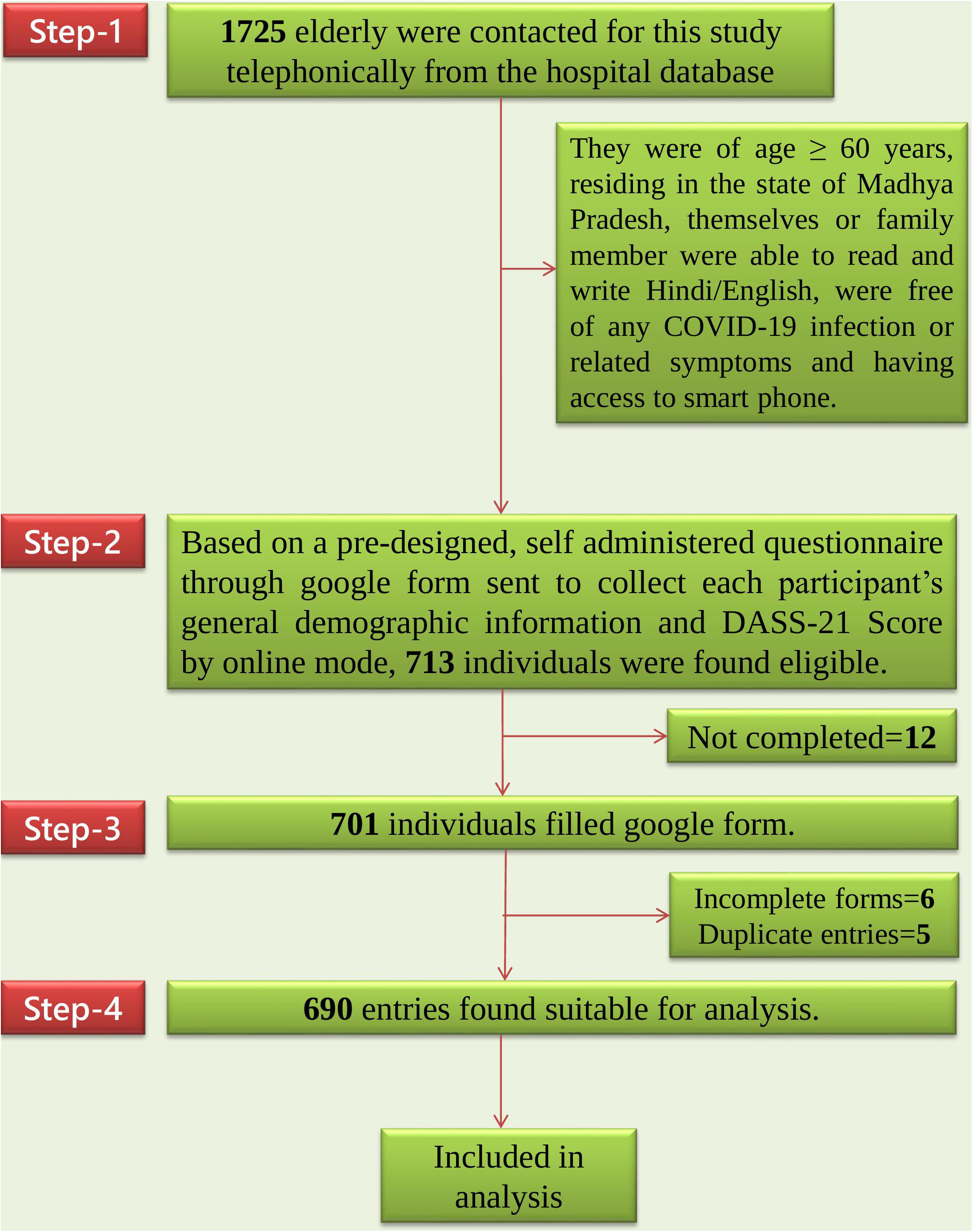
Flowchart showing Non-response rate at different steps.

**Figure 2:**
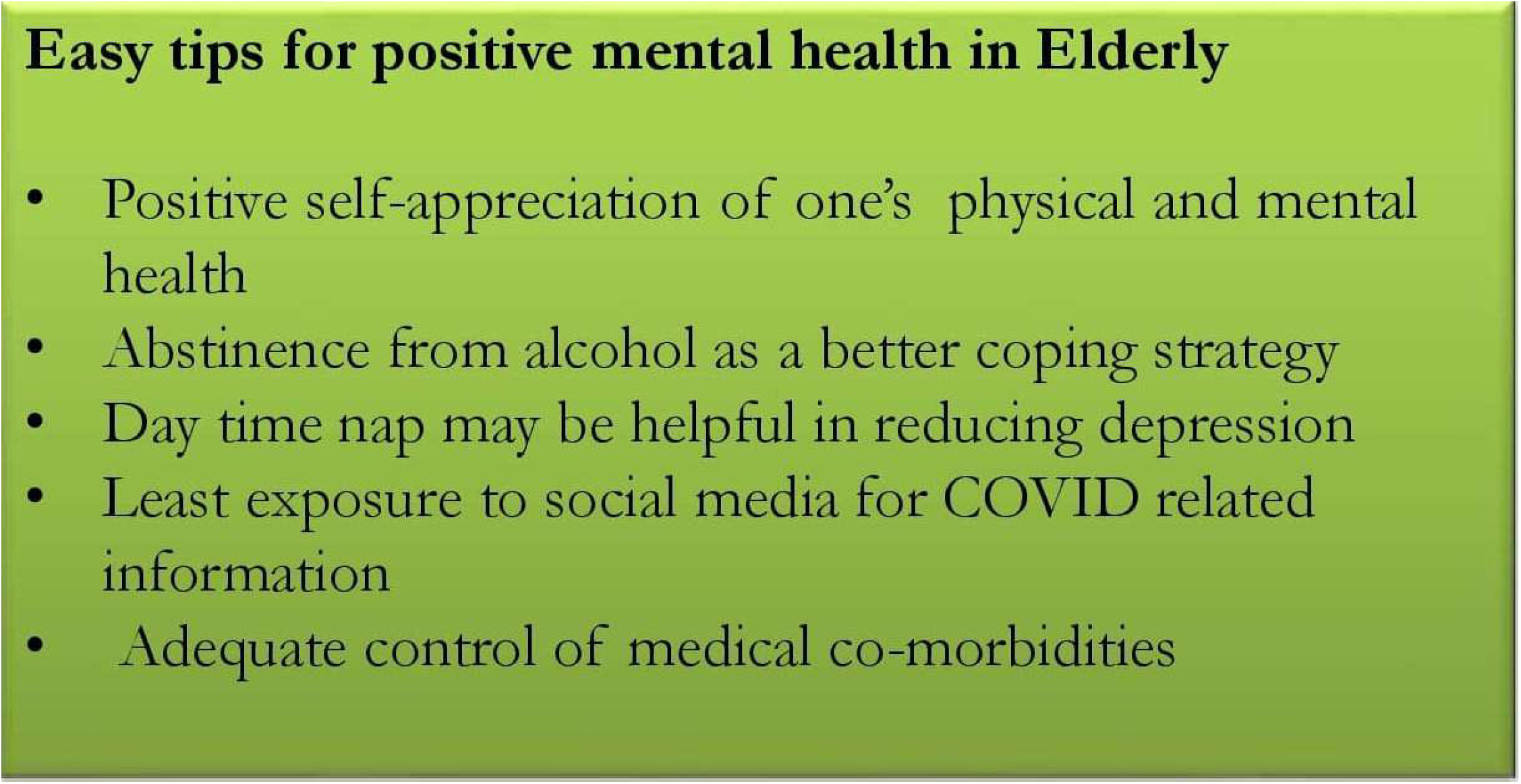
Easy tips for positive mental health in Elderly.

## Notes

### Competing Interest Statement

The authors have declared no competing interest.

### Funding Statement

Department of Science and Technology (DST), SATYAM (Science and Technology of Yoga and Meditation), Government of India for Financial Support vide sanction order reference number DST/SATYAM/COVID-19/2020/137

### Author Declarations

Approval of Institutional Human Ethics Committee of All India Institute of Medical Sciences, Bhopal, Madhya Pradesh, India (Letter No. IHEC-LOP/2020/EF0221, dated: 24/12/2020).

## REFERENCES

1. World Health Organization (WHO). (2020a, March 11). WHO Director-General’s opening remarks at the media briefing on COVID-19. Retrieved March, 2020, from https://www.who.int/dg/speeches/detail/who-director-general-s-opening-remarks-at-themedia-briefing-on-covid-19

2. Xiang YT, Yang Y, Li W, et al. Timely mental health care for the 2019 novel coronavirus outbreak is urgently needed. LANCET PSYCHIAT. 2020;7(3):228–9 doi: 10.1016/S2215-0366(20)30046-8

3. Liu S, Yang L, Zhang C. Online mental health services in China during the COVID-19 outbreak. LANCET PSYCHIAT 2020:7, e17–e18 doi: 10.1016/S2215-0366(20)30077-8

4. Wheaton MG, Abramowitz JS, Berman NC, et al. Psychological predictors of anxiety in response to the H1N1 (swine flu) pandemic. Cogn. Ther. Res. 2012;36(3):210–8 doi: 10.1007/s10608-011-9353-3

5. Wu P, Fang Y, Guan Z, et al. The psychological impact of the SARS epidemic on hospital employees in China: exposure, risk perception, and altruistic acceptance of risk. Can J Psychiatry. 2009;54(5):302–11 doi:10.1177/070674370905400504

6. Lee SA. Coronavirus Anxiety Scale: A brief mental health screener for COVID-19 related anxiety. Death Stud. 2020;44(7):393–401. doi: 10.1080/07481187.2020.1748481.

7. Zhang Y, Ma ZF. Impact of the COVID-19 pandemic on mental health and quality of life among local residents in Liaoning Province, China: A cross-sectional study. Int. J. Environ. Res. Public Health. 2020;17(7):2381 doi: 10.3390/ijerph17072381.

8. Wang C, Pan R, Wan X, et al. Immediate psychological responses and associated factors during the initial stage of the 2019 coronavirus disease (COVID-19) epidemic among the general population in China. Int. J. Environ. Res. Public Health. 2020;17(5):1729 doi: 10.3390/ijerph17051729

9. Gao J, Zheng P, Jia Y, Chen H, Mao Y, Chen S, Wang Y, Fu H, Dai J. Mental health problems and social media exposure during COVID-19 outbreak. PLoS One. 2020;15(4):e0231924 doi:10.1371/journal.pone.0231924

10. Wu P, Fang Y, Guan Z, et al. The psychological impact of the SARS epidemic on hospital employees in China: exposure, risk perception, and altruistic acceptance of risk. Can J Psychiatry. 2009;54(5):302–11 doi: 10.1177/070674370905400504.

11. Roy D, Tripathy S, Kar SK, et al. Study of knowledge, attitude, anxiety & perceived mental healthcare need in Indian population during COVID-19 pandemic. Asian j psychiatr. 2020;1;51:102083 doi: 10.1016/j.ajp.2020.102083

12. Gorrochategi MP, Munitis AE, Santamaria MD, et al. Stress, anxiety, and depression in people aged over 60 in the COVID-19 outbreak in a sample collected in Northern Spain. Am J Geriatr Psychiatry. 2020;1;28(9):993–8 doi: 10.1016/j.jagp.2020.05.022.

13. Sepúlveda-Loyola W, Rodríguez-Sánchez I, Pérez-Rodríguez P, et al. Impact of social isolation due to COVID-19 on health in older people: mental and physical effects and recommendations. J. Nutr. Health Aging. 2020;24(9):938–47 doi: 10.1007/s12603-020-1500-7

14. Armitage R, Nellums LB. COVID-19 and the consequences of isolating the elderly. Lancet Public Health, 2020;5:5e256 doi: 10.1016/S2468-2667(20)30061-X

15. Viswanath B, Maroky AS, Math SB, et al. Psychological impact of the tsunami on elderly survivors. Am J Geriatr Psychiatry. 2012;20(5):402–7 doi: 10.1097/JGP.0b013e318246b7e9

16. Choi KW, Chen CY, Stein MB, et al. Assessment of bidirectional relationships between physical activity and depression among adults: A 2-sample mendelian randomization study. JAMA Psychiatry 2019;76:399–408 doi: 10.1001/jamapsychiatry.2018.4175

17. Wang C, Pan R, Wan X, et al. Immediate Psychological Responses and Associated Factors during the Initial Stage of the 2019 Coronavirus Disease (COVID-19) Epidemic among the General Population in China. Int J Environ Res Public Health. 2020;17(5):1729 doi: 10.3390/ijerph17051729

18. Hwang TJ, Rabheru K, Peisah C, Reichman W, Ikeda M. Loneliness and social isolation during the COVID-19 pandemic. Int Psychogeriatr. 2020;32(10):1217–1220 doi: 10.1017/S1041610220000988

19. Djernes JK. Prevalence and predictors of depression in populations of elderly: A review. Acta Psychiatr. Scand., 2006;113(5):372–87 doi: 10.1111/j.1600-0447.2006.00770.x.

20. Byers AL, Yaffe K, Covinsky KE, et al. High occurrence of mood and anxiety disorders among older adults: The National Comorbidity Survey Replication. Arch. Gen. Psychiatry. 2010;67(5):489–96 doi: 10.1001/archgenpsychiatry.2010.35.

21. Usher K, Jackson D, Durkin J, et al. Pandemic-related behaviours and psychological outcomes; A rapid literature review to explain COVID-19 behaviours. Int. J. Ment. Health Nurs. 2020;29(6):1018–34 doi: 10.1111/inm.12790

22. Faraut B, Nakib S, Drogou C, et al. Napping reverses the salivary interleukin-6 and urinary norepinephrine changes induced by sleep restriction. J Clin Endocrinol Metab. 2015;100:E416–26 doi: 10.1210/jc.2014-2566.

23. Faraut B, Boudjeltia KZ, Dyzma M, et al. Benefits of napping and an extended duration of recovery sleep on alertness and immune cells after acute sleep restriction. Brain Behav Immun. 2011;25:16–24 doi: 10.1016/j.bbi.2010.08.001

24. Brooks A, Lack L. A brief afternoon nap following nocturnal sleep restriction: which nap duration is most recuperative? Sleep. 2006;29:831–40 doi: 10.1093/sleep/29.6.831

25. Baran B, Mantua J, Spencer RM. Age-related changes in the sleep-dependent reorganization of declarative memories. J. Cogn. Neurosci. 2016;28(6):792–802 doi: 10.1162/jocn_a_00938

26. Antonenko D, Diekelmann S, Olsen C, et al. Napping to renew learning capacity: enhanced encoding after stimulation of sleep slow oscillations. Eur. J. Neurosci. 2013;37(7):1142–51 doi: 10.1111/ejn.12118

27. Yang Y, Li W, Zhang Q, et al. Mental health services for older adults in China during the COVID-19 outbreak. LANCET PSYCHIAT. 2020;7(4):e19 doi: 10.1016/S2215-0366(20)30079-1

28. Yeung DY, Fung HH. Age differences in coping and emotional responses toward SARS: a longitudinal study of Hong Kong Chinese. Aging Ment Health. 2007;11(5):579–87 doi: 10.1080/13607860601086355.

29. Pal S, Kalra S, Jain S. COVID-19 Pandemic: Mental Health and Coping Strategies. J. Clin. Diagnostic Res. 2020;14(12) doi: 10.1016/j.jad.2020.08.001

